# Adolescents’ knowledge of pain medication – can a specific teaching program in primary school improve knowledge and attitudes toward the use of pain medication?

**DOI:** 10.1101/2023.05.27.23290626

**Authors:** Alessandro Andreucci, Anne Estrup Olesen, Camilla Merrild, Heidur Hardardóttir, Nabil Al-Janabi, Malene Kjær Bruun, Rikke Møller Larsen, Michael Skovdal Rathleff

**Affiliations:** Center for General Practice at Aalborg University, Aalborg University, Aalborg, Denmark; Department of Clinical Pharmacology, Aalborg University Hospital, Aalborg, Denmark; Department of Clinical Medicine, Aalborg University, Aalborg, Denmark; School administration, Aalborg Municipality; Department of Health Science and Technology, Faculty of Medicine, Aalborg University, Aalborg, Denmark

## Abstract

**Objectives:** The aim of this study was to investigate how a specifically designed teaching program for adolescents on the subject of “pain medication” affects their knowledge and attitudes regarding pain medication.

**Methods:** This prospective interventional study used both quantitative and qualitative methods. The teaching intervention was co-developed with end-users. Adolescents completed a questionnaire at 3 time points: 1) at baseline before the teaching intervention, 2) immediately after the intervention and 3) at follow-up after approximately 1-2 months, depending on the schools’ availability. A qualitative component with interviews on a subsample of participants was carried out between baseline and the 1-2 months follow-up.

**Results:** Nine classes, corresponding to 181 adolescents with a median age of 14 were exposed to the teaching intervention. 22% used pain medication at least once a week at baseline. Their baseline knowledge regarding the mechanism of action, side effects, dosage, and alternative methods to treat pain was low. Their levels of knowledge improved after the teaching intervention and we observed higher levels of knowledge and less uncertainty. However, despite the immediate positive effect, the retainment of knowledge was slightly reduced at the 1–2-month follow-up.

**Discussion:** Our intervention increased the overall knowledge on pain medication and reduced the adolescents’ uncertainty. However, the retainment of knowledge was reduced after 1–2-month follow-up. Future interventions carried out on a longer time-span and with the inclusion of parents and delivery of online material might be designed to improve retainment of knowledge.

## Introduction

Chronic musculoskeletal (MSK) pain affects up to 40% of children and adolescents (King et al., 2011). Living with chronic MSK pain issues a major burden on both adolescents and their families (Nielsen et al., 2014; Palermo, 2000). There are many different treatments for pain conditions, but first-line management often include pain medications (Caes et al., 2018). Within the last 30 years, pain medication use among adolescents has more than doubled, especially the use of over-the-counter (OTC) pain medication (paracetamol and NSAID’s) (Nielsen et al., 2014).

A recent systematic review showed that up to 42% of children and adolescents in school settings use pain medication on a regular basis (Al-janabi et al., 2021). Adolescents may start using OTC pain medication after initial advice from their parents, general practitioner, pharmacist, friends, teachers, sports coach, or media (TV, internet, newspapers) (Gualano et al., 2015; Jensen et al., 2014; Shehnaz et al., 2014; Skarstein et al., 2019). Despite the commonality of using pain medication, adolescents have insufficient knowledge of how pain medication work, risk of side effects and its effectiveness on their pain complaint (Hämeen-Anttila and Bush, 2008; Shehnaz et al., 2014; Stoelben et al., 2000; Wilson et al., 2010). This lack of knowledge may put adolescents at risk of using pain medication inappropriately without consideration for potential adverse events (Skarstein et al., 2018).

Adolescence is a stage of life where individuals form their identity and become increasingly independent in the development of attitudes and behaviours (Skarstein et al., 2018). The behaviours regarding use of pain medication as a coping strategy are formed during adolescence and can be maintained during adulthood (Andersen et al., 2009; Jonassen et al., 2021; Skarstein et al., 2018). This may become a problem if pain medication is used in the wrong way and with the wrong indications. Despite OTC pain medication is easily available there are documented side effects of long-term use of OTC pain medication, including both minor (headache and abdominal pain) and major side effects (gastrointestinal bleeding, reduced cardiovascular health and renal failure).

Evidence from other research areas like tobacco use, drugs and alcohol show that teaching interventions can improve the knowledge among adolescents (McBride et al., 2004; Thomas et al., 2013). This is particularly relevant in the field of pain medication as previous research indicates that insufficient knowledge about rational use, is one of the factors driving the use of pain medication among adolescents (Hämeen-Anttila and Bush, 2008; Shehnaz et al., 2014; Stoelben et al., 2000; Wilson et al., 2010). Therefore, this study aimed to investigate how a specifically designed teaching program for adolescents on the subject of “pain medication” affects their knowledge and attitudes regarding pain medication. The working hypothesis was that a teaching intervention can be used to improve the knowledge and attitudes towards the use of pain medication in adolescents.

## Methods

### Design

This was a prospective interventional study using quantitative and qualitative methods. This study included end-users from the beginning in the co-creation of both research question, design, questionnaires, and development of the teaching materials and its implementation. Adolescents completed a questionnaire at 3 time points: 1) at baseline before the teaching, 2) immediately after the teaching and 3) at follow-up after approximately 1-2 months, depending on the schools’ availability. A qualitative component with interviews on a subsample of participants was carried out between baseline and the 1-2 months follow-up. The study aligned with the GPDR and privacy regulations regarding the collection of data from participants below 18 years of age.

### Setting and participants

RML (consultant at Aalborg Municipality) reached out to several schools in Aalborg and invited to take part to the project. Three schools located in different socioeconomic areas in Aalborg, Denmark, accepted to participate. Adolescents at the 6th and 9th grade (age 12-15 years old) from these schools were recruited. Adolescents were provided oral information about the study on the day of baseline data collection. The adolescents’ parents were informed about the study prior to the date of data collection by the school coordinators through the software AULA (Fællesoffentlige Brugerportalsinitiativ (BPI), Denmark), which is a communication platform for employees, parents and students used in the primary schools and in day care in Denmark. Through this platform, parents had the opportunity to object towards their child’s participation in the study. Participation was voluntary and completely anonymous.

### Procedure

This study included specific stages regarding the development of questionnaire and teaching material, the data collection and the lecture on proper use of pain medication, as outlined in figure 1.

**Figure 1.**
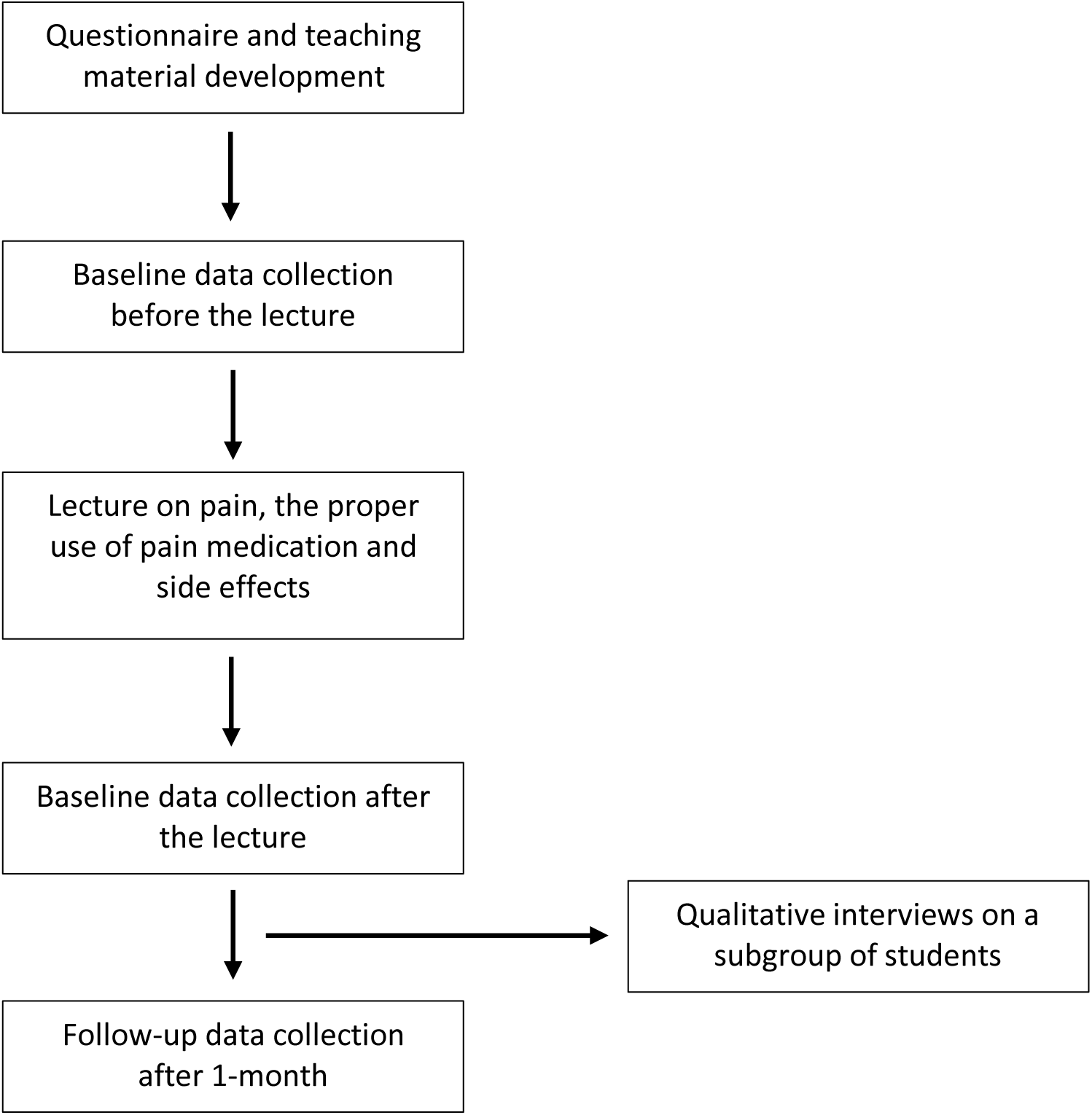
Stages of the study

### Intervention

#### Intervention development

The teaching intervention was designed through a co-creation process with the “SSP-Trivsel” section at Aalborg Municipality. This unit is specialised in delivering education on topics such as alcohol, smoking and drugs use. Initially researchers and the school coordinator from the municipality discussed the learning goals, time availability and how teaching should support reaching these learning goals. This information was then used by two university professors (AEO and MSR) who made the first version of the teaching materials and the different engagement methods that would be used during the teaching. This was then revised after input from RML and two adolescents. The intervention included a specifically designed lecture focused on topics such as the proper use of pain medication for different pain conditions, taking the medication at the recommended therapeutic dose and in accordance with leaflet instructions, the mechanism of action, the efficacy of pain medication and potential side-effects associated with use. It was based on both theories delivered by lectures, discussions, use of artifacts (e.g., cases of pain medication).

#### Intervention delivery

The lecture was delivered by one of two young medical doctors wearing white coats, who were assisted by a member of the research team (A.A.) during the lecture delivery and data collection. The lecture was delivered with the support of a power-point presentation and of a body mannequin to outline the human body and the mechanisms of action of the different types of pain medication (e.g., oral vs topical). Adolescents were also handed the pain medication packages to provide them with the opportunity to read the medication leaflet and become more familiar with the correct mode of use of pain medication, dosage and potential side-effects.

### Questionnaire

The questionnaires were developed in collaboration with RML to ensure that it would cover all the domains needed to achieve the learning goals of the project. To ensure the face, construct and content validity of the questionnaire, an initial version was drafted and pilot-tested several times with volunteer adolescents of the same age range of participants included in the project. Following feedback from adolescents, the questionnaire was implemented and a final version to use in the data collection stage was obtained. The questionnaire included the following domains: 1) Demographics; 2) Use of pain medication; 3) Pain and pain management; 4) Side-effects and harms related to use; 5) Dose and overdoses; 6) Medication appearance; 7) Mechanism of action, allergies and differences in use between adults and children/adolescents; 8) Attitudes towards use of pain medication, reasons for use and social context.

The questionnaire used in the data collection immediately after the lecture was a shortened version of the baseline questionnaire. There were small modifications regarding some questions, as outlined in appendix 1. The questionnaire used at the 1-2 month follow-up was the same used at baseline, and the only modification regarded the time period for the frequency of pain medication use (appendix 1). Questionnaires were filled on tablets, computers and smartphones via an electronic data capture tool (REDCap) (Harris et al., 2009).

### Interviews and interview guide

Semi-structured interviews were performed by a member of the research team (M.K.B.) to explore more in-depth themes on the use and knowledge and attitudes towards the use of pain medication. Interviews lasted on average 25 minutes and were conducted via Microsoft Teams.

An interview guide (appendix 2) was developed by the main author of this paper with the support of a research member with expertise in qualitative research (C.M.) and pilot-tested with volunteer adolescents to ensure comprehensibility of the questions.

The interview guide included open-ended questions to explore the following domains: 1) pain management; 2) tendency to use pain medication; 3) pain medication knowledge in terms of dosage, side effects and appearance; 4) general attitude regarding the use of pain medication; 5) use of pain medication as a preventive strategy; 6) use of pain medication for daily activities; 7) bringing pain medication to school / physical activity.

### Data analysis

#### Quantitative analysis

Descriptive analysis of the sample using data collected at baseline (before and after the lecture) and at 1–2-month follow-up were carried out. Data are shown as means and standard deviations (SD) or as percentages (%) where appropriate. Analysis of the change in use and knowledge and attitudes towards the use of pain medications were performed, with comparison between data collected at baseline before the teaching, after the teaching and at the 1–2-month follow-up. The change in uncertainty was calculated by comparing the proportion of adolescents who replied “I don’t know” at the different time-points. Proportion of estimates with 95% confidence intervals were shown as forest plot.

#### Qualitative analysis

The qualitative data was coded thematically, to explore more in-depth themes on the use and knowledge and attitudes towards the use of pain medications, all interviews were recorded and transcribed verbatim, and two authors coded and thematized independently of each other before comparing notes and agreeing on the final themes (Rivas, 2012).

## Results

### Sample description

Three schools for a total of 9 classes participated to the study. Overall, 181 adolescents completed the questionnaire at baseline before the lecture, of which 174 completed the questionnaire after the lecture. One-hundred-sixty-eight adolescents provided data at the 1–2-month follow-up.

Table 1 outlines the baseline characteristics of the sample.

**Table 1.** Baseline sample characteristics.

| **Table 1. Baseline sample characteristics** |  |  |
| --- | --- | --- |
|  | Median | Interquartile range |
| Age | 14 | 13 - 15 |
|  | N | % |
| Gender |  |  |
| *Boys* | 91 | 50.3 |
| *Girls* | 88 | 48.6 |
| *Other* | 2 | 1.1 |
| Ethnicity |  |  |
| *Danish* | 167 | 92.3 |
| *Non-Danish* | 14 | 7.7 |

### Self-report use of pain medication

At baseline before the lecture, adolescents reported that the most used pain medication was paracetamol, followed by ibuprofen and medication in form of gel/creme (table 2). In addition, adolescents answered “Yes” or “Sometimes” (baseline: n=137, 76%) to the question “Do you take pain medication when you have pain in your body?”, and were subsequently asked further questions regarding the frequency and mode of use (table 2). Approximately 22% of adolescents reported using pain medication at least once a week at baseline. Regarding the mode of use, 63.5% reported using pain medication a single day each time and 32.1% several days each time.

**Table 2.**
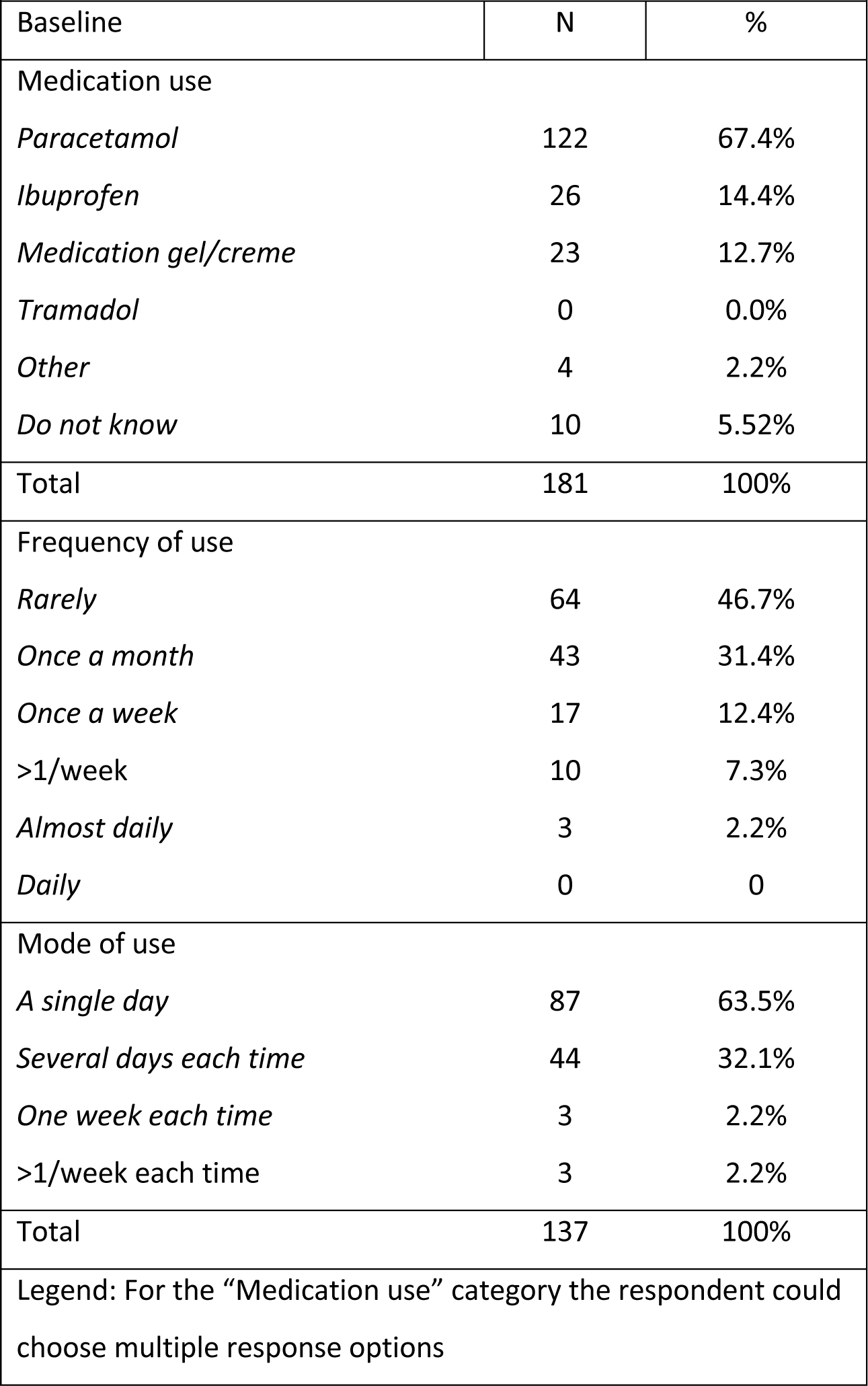
Frequency of use of pain medication and mode of use.

### Attitudes towards use of pain medication, reasons for use and social context

Before the lecture, adolescents reported that the main reasons for using pain medication were pain severity and duration, followed by the ability to sleep and to go to school (Table 3). One in every three adolescents reported to have been worried that taking pain medication could be harmful and 9.9% reported being worried of becoming addicted to pain medication. Twenty-eight percent of adolescents indicated “Yes” and 46% “sometimes” when asked about taking pain medication instead of going to their GP. Only, 26% indicated reading the leaflet when taking pain medication and 39% read it “sometimes”. Most commonly the advice for taking pain medication would come from the parents (84%), followed by the doctor (34%) and the adolescents’ own decision to take pain medication (33%).

**Table 3.** Attitudes towards use, reasons for use of pain medication and social context.

|  |  |  |  |  |  |
| --- | --- | --- | --- | --- | --- |
|  | N | % |  | N | % |
| Reasons for use |  |  | Social meaning use |  |  |
| *Pain frequency* | 52 | 28.7% | *It is cool* | 4 | 2.2% |
| *Pain severity* | 102 | 56.4% | *It is strange* | 1 | 0.6% |
| *Pain duration* | 101 | 55.8% | *One is weak* | 4 | 2.2% |
| *To prevent pain* | 12 | 6.6% | *It does not matter* | 100 | 55.3% |
| *To practice sport* | 26 | 14.4% | *It is embarrassing* | 0 | 0.0% |
| *To go to school* | 43 | 23.8% | *Other* | 15 | 8.3% |
| *To meet friends* | 14 | 7.7% | *None of the above* | 57 | 31.5% |
| *Low mood* | 3 | 1.7% | Bringing pain medication to school |  |  |
| *Impaired sleep* | 75 | 41.4% | *No* | 117 | 64.6% |
| *Other* | 6 | 3.3% | *Yes* | 36 | 19.9% |
| *Do not use* | 18 | 9.9% | *Sometimes* | 28 | 15.5% |
| Use to avoid doctor |  |  | Bringing pain medication to sport training |  |  |
| *No* | 46 | 25.6% | *No* | 139 | 76.8% |
| *Yes* | 51 | 28.3% | *Yes* | 23 | 12.7% |
| *Sometimes* | 83 | 46.1% | *Sometimes* | 19 | 10.5% |
| Reading the leaflet |  |  | When to use pain medication |  |  |
| *No* | 63 | 34.8% | *As soon as I feel pain* | 6 | 3.3% |
| *Yes* | 47 | 26.0% | *Only if it is very bad pain* | 71 | 39.2% |
| *Sometimes* | 71 | 39.2% | *When pain affects ability to do things* | 102 | 56.4% |
| Fear of effect (yes) | 60 | 33.1% | *Other* | 2 | 1.1% |
| Addiction fear (yes) | 18 | 9.9% | Borrow pain medication from friends (Yes) | 32 | 17.7% |
| Total | 181 | 100% | Total | 181 | 100% |
| Legend: For the reasons for use the respondent could choose multiple response options |  |  |  |  |  |

### Knowledge of pain medication

The most known pain medication at baseline was paracetamol, followed by ibuprofen and medication in form of gel/creme (table 4). At the 1–2-month follow-up, the most known pain medication was still paracetamol, followed by medication in form of gel/crème and ibuprofen.

**Table 4.** Knowledge of pain medication.

| **Table 4. Knowledge of pain medication** |  |  |  |  |  |
| --- | --- | --- | --- | --- | --- |
| Baseline | N | % | 1–2-month follow-up | N | % |
| Medication knowledge |  |  | Medication knowledge |  |  |
| *Paracetamol* | 130 | 71.8% | *Paracetamol* | 125 | 74.4% |
| *Ibuprofen* | 45 | 24.9% | *Ibuprofen* | 55 | 32.7% |
| *Medication gel/creme* | 50 | 27.6% | *Medication gel/creme* | 73 | 43.5% |
| *Tramadol* | 6 | 3.3% | *Tramadol* | 13 | 7.7% |
| *None of them* | 6 | 3.3% | *None of them* | 3 | 1.8% |
| Total | 181 | 100% |  | 131 | 100% |
| Legend: For the “Medication knowledge” category the respondent could choose multiple response options |  |  |  |  |  |

### Knowledge of pain management

Before the lecture 69% of adolescents replied that pain medication is not the only method to treat pain, 46% indicated that they knew alternative methods to treat pain and 90% that pain can be treated without pain medication. These figures increased after the lecture and decreased at the 1-2 month follow-up, despite being still higher than before the lecture (Figure 1).

**Figure 1.**
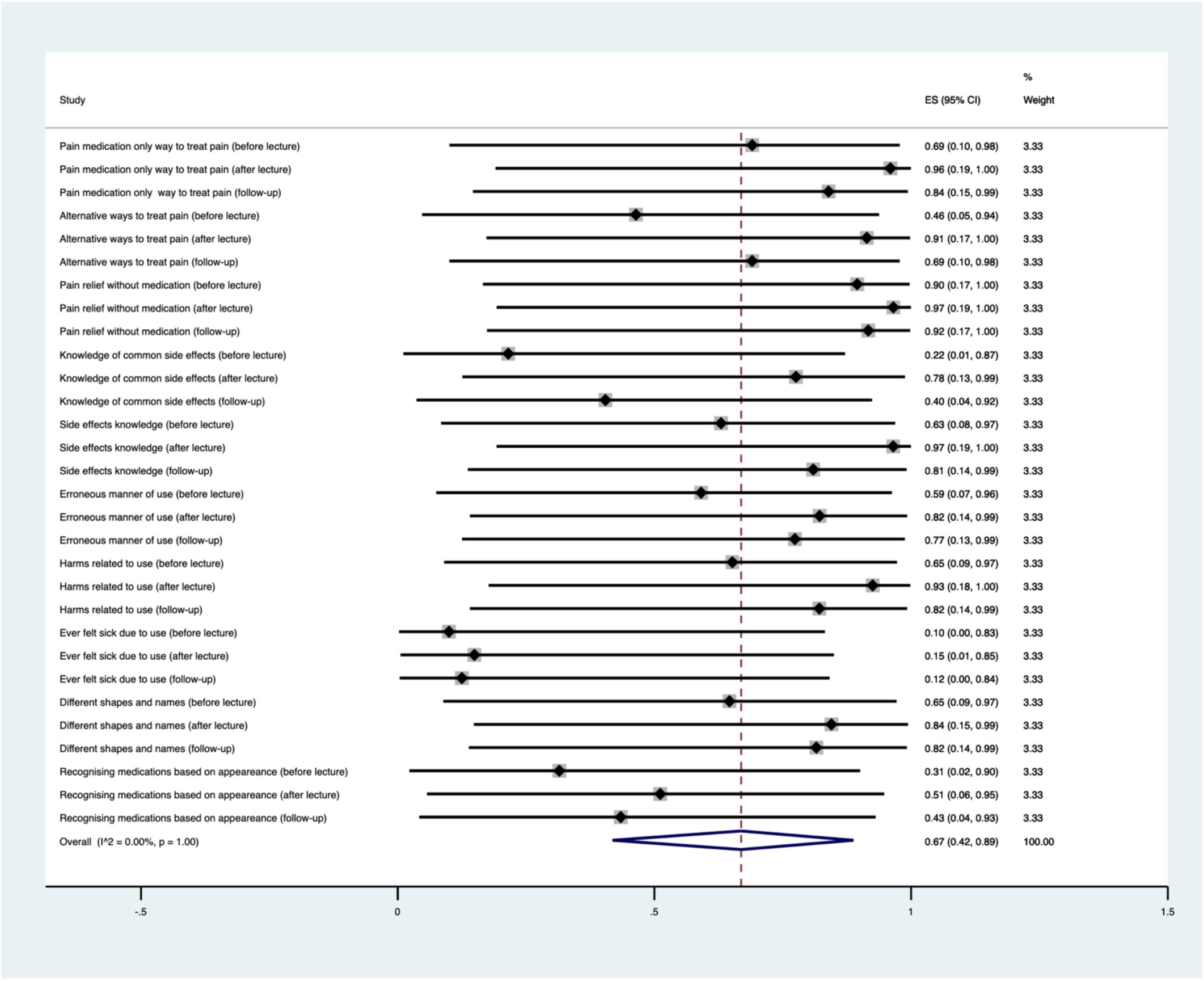
Knowledge of pain, pain management, side effects, medication mode of use and appearance

Across all questions there was an overall 16% decrease in uncertainty after the lecture which changes to 10.3% after 1-2 months (range 5.9-14.3%).

### Side-effects and harms related to use

Before the lecture, 63% of adolescents indicated that pain medication can have side effects and 22% knew that using pain medication could result in common side effects such as stomach problems, feeling tired or sleepy. The main source of knowledge of side effects was the family and friends (30.2%), followed by the general knowledge (20.8%) and the leaflet (16.3%), the doctor (12.9%), the internet (11.4%) and the pharmacist (6.4%) to a minor extent. Regarding the mode of use, 59% replied that pain medication can be taken in an erroneous manner, 65% that taking pain medication can be harmful and 10% had ever been sick following pain medication intake. These figures increased after the lecture and slightly decreased at the 1-2 month follow-up. (Figure 1).

There was an average overall 23% uncertainty decrease (from 32.9% before the lecture to 9.2% after the lecture). After 1-2 months, the average uncertainty was 16.7% (range 12.5-21.4%).

### Medication appearance

Before the lecture, 65% reported that the same medication can have more than one form and different names and 31% replied that they can recognize pain medication based on their appearance (e.g., the package). These figures increased both after the lecture and at the 1-2 month follow-up. (Figure 1). Before the lecture, the percentage of those who were uncertain was 43.4% and decreased to 31.0% after the lecture and to 30.4% after 1-2 months, with an average overall 12-13% uncertainty decrease.

### Dose and overdoses

Before the lecture, 91% correctly reported that pain medication cannot be taken without limit during the day, 71% that it may be necessary to go to the doctor due to taking too much pain medication and 53% reported knowing the right dose of pain medication to take when in pain. When asked if taking too many vitamins, paracetamol and cough medicine could be harmful, 43%, 70% and 44% replied “yes”, respectively (Figure 2). These proportions increased after the lecture and slightly decreased at the 1-2 month follow-up. There was an average overall 15% uncertainty decrease from before (28.2%) to after the lecture (12.9%). After 1-2 months, the overall uncertainty was 17.4% (range 5.9-25.0%).

**Figure 2.**
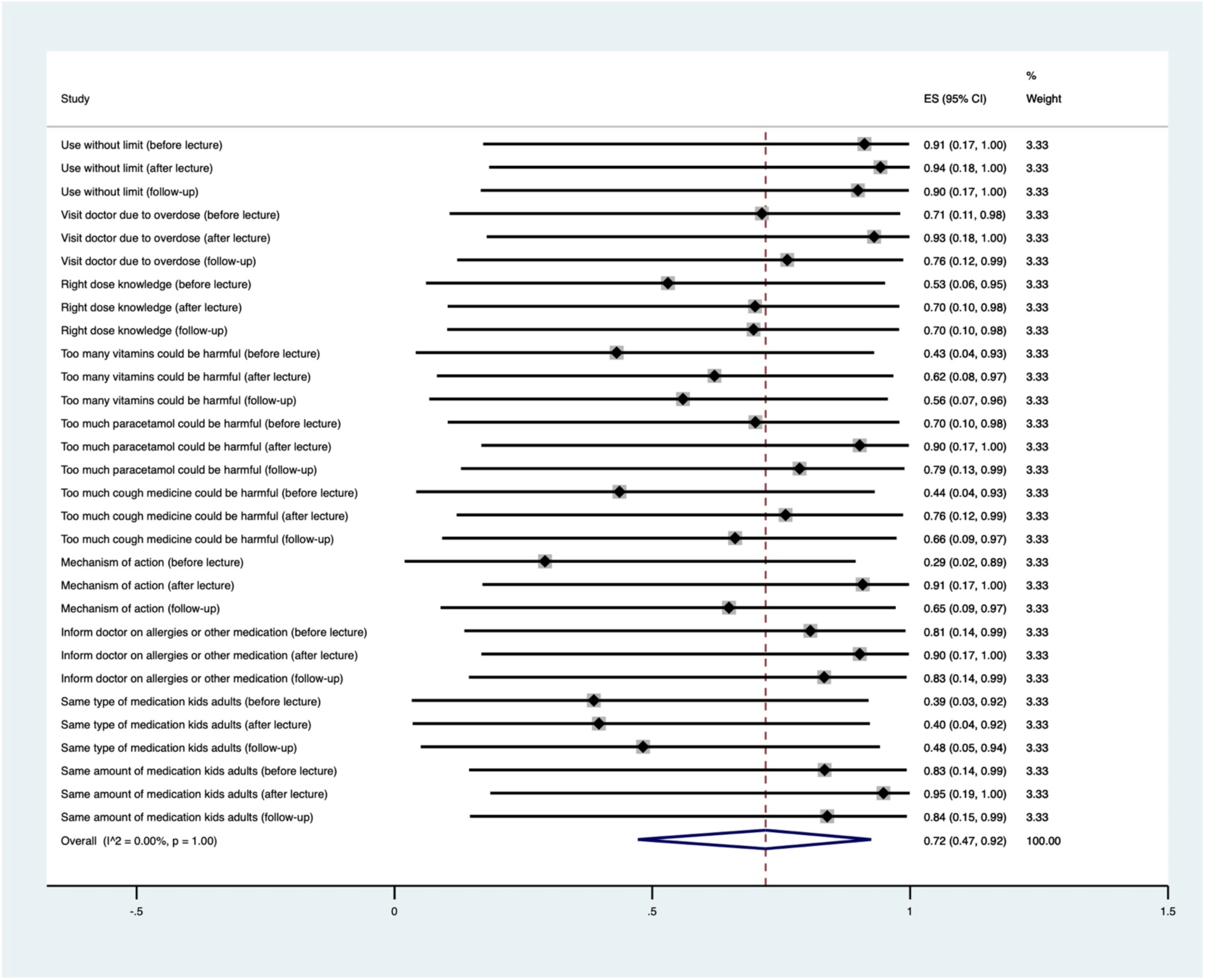
Knowledge of dosage and overdose, mechanisms of action, allergies and differences between adults and kids

### Mechanism of action, allergies and differences in use between adults and children/adolescents

Before the lecture, 29% of adolescents reported knowing how pain medication function in the body, 81% that they should inform the doctor if they are allergic to a medication or currently taking another medication, 39% reported that kids and adults can take the same type of pain medication and 83% that kids and adults cannot take the same amount of pain medication. These proportions increased after the lecture and decreased at the 1-2 month follow-up. (Figure 2).

There was an average overall 15% uncertainty decrease from before (20.4%) to after the lecture (4.9%). After 1-2 months, the overall uncertainty was 10.7% (range 10.1-11.3%).

### Interviews

Five adolescents participated in the semi-structured interviews and five overall themes emerged.

#### Use to cope with daily activities and general attitude regarding use

The general tendency was not to use pain medication as a first option. Generally, adolescents expressed a positive attitude regarding the use of pain medication when in pain (especially when experiencing severe pain), as long as the quantity was not excessive and the use not too frequent, as this might affect the efficacy in their opinion. In addition, the interviewees reported concerns regarding addiction and side effects.

#### Knowledge of pain medication, mechanism of action and dose

Adolescents were familiar with different pain medication classes and formulations, such as paracetamol/Ibuprophen, tablets/effervescent tablets/topical cream and gel and on when to use them (e.g., paracetamol for fever, topical administration for local pain). Adolescents reported that the advice regarding taking pain medication mainly came from family members (parents or grandparents) and the doctor.

Regarding side effects, adolescents were familiar with common side effects such as stomachache, nausea, diarrhea and the source of information was the parent, the medication leaflet, and our lecture. Adolescents were familiar with the general mechanisms of action and the difference in functioning between oral and topical administration and reported that their knowledge came from our lecture.

#### Use of pain medication to prevent pain and in the social context (family, sport, school)

Adolescents reported knowing someone (i.e., parents, relatives) that uses pain medication regularly following surgery and they believed that there was no problem associated with this use because it was prescribed by a doctor. However, they reported not talking about pain medication with their peers usually, because they are interested in other topics.

Generally, adolescents reported not using pain medication in relation to playing sport, unless in connection to injuries, and expressed concerns on becoming dependent on pain medication.

Similarly, pain medication at schools were used only sporadically and in connection with pain conditions (e.g., headache, sore throat) but not to improve school performances. However, they were not concerned regarding bringing pain medication to the sport training or at school if there is someone who can supervise the use.

## Discussion

### Main findings

A substantial proportion of school attending adolescents use pain medication at least once a week. They bring pain medication to school, sport and when they are with friends or in other leisure activities. Their baseline knowledge regarding the mechanism of action, side effects, dosage, and alternative methods to treat pain was low. After the lecture the level of knowledge changes and we observed higher levels of knowledge and less uncertainty. However, despite the immediate positive effect, the retainment of knowledge was slightly reduced at the 1–2-month follow-up.

### Comparison with previous studies

One in every four adolescents reported using pain medication at least once a week. This is in line with previous studies from Norway (Jonassen et al., 2021; Skarstein et al., 2014). A lack of alternative treatments may explain the high use due to desire to cope with tasks in daily life. (Lagerløv et al., 2016; Skarstein et al., 2018). The majority of adolescents in the study reported that pain medication should be used when pain affects the ability to do things or when pain is severe. This findings is consistent with previous findings (Lagerløv et al., 2016) and support the notion that some adolescents do indeed have a rational use of pain medication. The large differences in self-reported use-patterns may reflect different pain coping styles adopted by adolescents or that other causes than pain may affect the use pattern. (Garmy et al., 2019; Lagerløv et al., 2016).

One in every three adolescents reported not reading the medication leaflet when using pain medication. This is in contrast to previous findings of studies from Norway and the U.S. showing that most adolescents read the package information and found the information useful (Lagerløv et al., 2016; Nabors et al., 2004). This may originate from cultural differences and specific legislations of countries that can affect the availability of OTC pain medication and familiarity with their use, as observed in previous studies (Al-janabi et al., 2021; Shehnaz et al., 2014)

Despite most adolescents correctly reported that pain medication cannot be taken without any limit, only half of them reported knowing the right dose before the teaching intervention. This might be a consequence of the parents’ role in administering pain medication to their children (Skarstein et al., 2018). Pain medication diversion (i.e., borrowing and sharing prescribed or over-the counter pain medication to others) is a common behaviour among adolescents widely described in the literature (Hämeen-Anttila and Bush, 2008; Shehnaz et al., 2014; Skarstein et al., 2018). Figures from our sample are in agreement with the available evidence, as 18% adolescents at baseline and 27% at the 1-2 months follow-up reported ever borrowing pain medication from friends. In the interviews, adolescents reported that pain or pain management was not a topic usually discussed with their peers. This is in line with previous findings (Lagerløv et al., 2016). Recent evidence also shows that in addition to pain management there are other common reasons that can explain the adolescents’ use of pain medication. These include prevention of pain and tiredness, placebo for stress and anxiety control, sleep problems, easy access to pain medication that might promote a non-therapeutic use and peer pressure (Kiza et al., 2021; Skarstein et al., 2019).

### Strengths and limitation

A major strength of this study is the thorough process of development of teaching material in collaboration with Aalborg municipality and schools, which enabled to tailor the intervention to the adolescents’ cognitive and literacy level and ensured comprehensibility. In addition, this process allowed to integrate the learning outcomes into already defined learning outcomes for science teaching in primary school without the requirement of extra hours. This could have had a beneficial effect on the process of recruiting process schools for the study. Stakeholder involvement and integrated learning outcomes will also increase the chance of future implementation of our teaching program. Another strength is the use of both quantitative and qualitative methods to investigate the study objectives, which allowed to obtain quantitative data on the use, knowledge and attitudes towards the use of pain medication as well as more in-depth insights on the reasons and beliefs regarding these topics. We used a comprehensive pilot-tested questionnaire to collect data on use and knowledge of pain medication. This improves our confidence in the findings and reduce the risk of reporting bias (Delgado-Rodríguez and Llorca, 2004). In addition, the interview guide was pilot-tested and interviews were performed by a research student in physiotherapy to avoid a potential perception of authority (e.g., in case a doctor performed the interview) that could result in reduced openness and truthfulness regarding the answer provided (e.g., excessive or improper use of pain medication). Adolescents were recruited from schools from different socioeconomic areas in Aalborg, to limit differences in the use and knowledge of pain medication which might be associated to socioeconomic factors (Cantarero-Arévalo et al., 2014; Holstein et al., 2019). The long questionnaires used could also have resulted in “questionnaire tiredness” that might occur when replying to a long questionnaire and especially in adolescents who have a lower attention-span.

However based on the high number of completed responses we believe this limitation is minor. Social desirability bias might have occurred regarding the questions inquiring on the reasons for use of pain medication (Delgado-Rodríguez and Llorca, 2004). Future studies may wish to capture pain medication use prospectively and not rely on self-report use only.

### Implications

Previous studies on adolescents found that pain or pain management are seldomly a topic of discussion with parents or friends, despite adolescents would like more guidance on this topic (Lagerløv et al., 2016). As pain is common among adolescents (King et al., 2011), this underlines the importance of interventions targeted at increasing the adolescents’ knowledge on pain and pain management, including the proper use of pain medication. In our study we found a slight decrease in the retainment of knowledge and increased uncertainty in the replies after 1–2-months. Future interventions might be designed together with stakeholders (i.e., schools, municipality) and include shorter lectures delivered in in several sections over a longer follow-up period, possibly as a part of the school curricula, to increase retention of knowledge. The interventions might also include the parents’ participation, as they are usually the main source of provision of pain medication (Jonassen et al., 2021), and school nurses, as they can play a key role in the identification of school adolescents at high risk of misuse of pain medication (Skarstein et al., 2019). In addition, information might be delivered online by means of short videos on social media, as their use is widespread among adolescents nowadays (Skarstein et al., 2019).

## Conclusions

A high proportion of school attending adolescents use pain medication frequently, despite a low knowledge regarding the mechanism of action, proper dosage, side effects, potential harms related to erroenous use, and general pain management. Our intervention increased the overall knowledge on pain medication and reduced the adolescents’ uncertainty. However, the retainment of knowledge was reduced after 1–2-month follow-up. Future interventions carried out on a longer time-span and with the inclusion of parents and delivery of online material might be designed to improve retainment of knowledge.

## Data Availability

All data produced in the present study are available upon reasonable request to the authors

## Acknowledgments

The authors would like to thank the school coordinators and schoolteachers in the Aalborg Municipality for their help in the preparation of the study and support regarding the organization of the lectures and data collection.

